# Machine Learning for Prediction of Childhood Mental Health Problems in Social Care

**DOI:** 10.1101/2024.05.03.24306756

**Authors:** Ryan Crowley, Katherine Parkin, Emma Rocheteau, Efthalia Massou, Yasmin Friedmann, Ann John, Rachel Sippy, Pietro Liò, Anna Moore

## Abstract

**Background:** Rates of childhood mental health problems are increasing in the United Kingdom. Early identification of childhood mental health problems is challenging but critical to future psycho-social development of children, particularly those with social care contact. Clinical prediction tools could improve these early identification efforts.

**Aims:** Characterise a novel cohort of children in social care and develop and validate effective Machine Learning (ML) models for prediction of childhood mental health problems.

**Method:** We used linked, de-identified data from the Secure Anonymised Information Linkage (SAIL) Databank to create a cohort of 26,820 children in Wales, UK, receiving social care services. Integrating health, social care, and education data, we developed several ML models. We assessed the performance, interpretability, and fairness of these models.

**Results:** Risk factors strongly associated with childhood mental health problems included substance misuse, adoption disruption, and autism. The best-performing model, a Support Vector Machine (SVM) model, achieved an area under the receiver operating characteristic curve (AUROC) of 0.743, with 95% confidence intervals (CI) of 0.724-0.762. Assessments of algorithmic fairness showed potential biases within these models.

**Conclusion:** ML performance on this prediction task was promising but requires refinement before clinical implementation. Given its size and diverse data, the SAIL Databank is an important childhood mental health database for future work.

## Introduction

### Childhood Mental Health Problems

The incidence and prevalence of childhood mental health problems are increasing in the UK, with a recent report placing the prevalence at approximately 16% (1). This increase likely stems from a confluence of factors including the COVID-19 pandemic, widening income inequality, social media usage, and increased pressure within school settings (2). Children in social care settings have a greater risk of poor mental health outcomes due to higher levels of Adverse Childhood Experiences (ACEs) and barriers to accessing care (3). Identifying childhood mental health problems is difficult because normal development and early symptoms of a disorder can be challenging to disentangle, children experience different symptoms as they age, and children may struggle to explain their feelings and behaviours (4). Identification for children with social care contact can be particularly difficult because ACEs can negatively impact development, and the care systems normally responsible for identifying problems in children (e.g. carers, GPs, and schools) are inconsistent and disrupted. Estimates on the rates of mental health problems in children in social care settings vary, with figures ranging between 19% and 38% (5,6). Despite the importance of early detection of mental health problems to facilitate provision of appropriate support, children with social care contact struggle to access assessment and subsequent treatment. This perpetuates difficulties as children’s early experience with psychopathology can lead to negative outcomes that affect them throughout adolescence and adulthood (7,8). It is therefore imperative to develop alternative solutions to support early identification of problems for this vulnerable group.

### Clinical Prediction Tools in Psychiatry

Despite the increasing burden of mental health problems on healthcare systems, growth in the number of mental health professionals is significantly outpaced by those afflicted (9). Clinical prediction tools can potentially improve outcomes and reduce resource burdens by identifying mental health problems early and guiding individuals towards appropriate support. Nevertheless, despite extensive research and promise of predictive risk tools, no machine learning (ML) models are clinically available for prediction of mental health in children (10).

Discrepancies between the vast potential for ML applications and corresponding lack of improvement in patient outcomes has been dubbed the “Artificial Intelligence (AI) chasm.” Low quality evaluations of model performance are common and an important cause of the chasm; evaluations are typically conducted via internal validation, without proper safeguards, and using methods that may overestimate performance (11). These issues are often magnified within psychiatry where models often suffer from low generalisability as assessments are predominantly conducted in homogeneous populations in affluent countries, datasets are typically smaller, and external validations of model performance are exceedingly rare (12–14). Using population-based, representative datasets can mitigate these limitations.

### SAIL Databank

The Secure Anonymised Information Linkage (SAIL) Databank is a national data safe haven providing approved researchers with linkable de-identified health, social care, and education datasets relating to the Welsh population (15). The Adolescent Mental Health Data Platform (ADP) contains data relating to children and young people aged 0-17 years and includes routinely collected data on demographics, education (e.g. attendance and attainment), health (e.g. outpatient care) and social care contact (e.g. child protection records). These datasets contain various mental health risk factors that can be used for model building. For social care, Children In Need Wales (CINW) was succeeded by Children Receiving Care and Support (CRCS) following the enactment of the Social Services and Well-being (Wales) Act in April 2016. Both datasets utilise ‘need for care and support’ as the all-encompassing indication for inclusion of children within the dataset and employ annual census collection methods that differ slightly in implementation. See Lee and colleagues (2022) for details of these databases (16).

### Study Aims

Overall, we strive to develop and analyse prototype ML models for the prediction of mental health problems in children under social care services using the SAIL databank. Since AI algorithms can reinforce historical patterns of systemic bias (17), we take an approach that integrates clinician perspectives, focuses on model interpretability, and assesses algorithmic fairness.

## Method

### Data

With support from ADP, we linked 18 datasets from SAIL (Supplemental Table 1). This linking process utilised demographic information and local identifiers to connect individuals to a unique Anonymous Linkage Field (ALF) identifier. Individuals were eligible for inclusion if they were age 10-17 years within the years 2013-2020 and had received social care at any time (e.g. appeared in either CINW or CRCS). Individuals were excluded if they were under 10 years old, could not be linked to the other datasets, or information on their mental health status was not available. All retrospective and subsequent data relating to these young people were included for analysis. The dataset was randomly split into a training set (70%), validation set (15%), and test set (15%).

### Mental Health Outcomes

Data collected on mental health events by the CINW/CRCS censuses were utilised for measurement of the outcome. As defined by CINW/CRCS, a child has a mental health problem if they are 10 years or older and meet any of the following criteria: have been diagnosed by a medical practitioner; have received Child and Adolescent Mental Health Services (CAMHS); or are on a waiting list for CAMHS. Mental health problems include depression, anxiety, eating disorders, self-harm, and other disorders but exclude substance misuse, autistic spectrum disorders, and other learning disabilities unless accompanied by mental health problems.

### Diagnosis/Intervention Codes

The richest clinical information in SAIL is found within diagnosis and intervention codes. Diagnosis codes within SAIL follow the International Classification of Disease, version ten (ICD-10) format. Intervention codes within SAIL follow the format of the Office of Population Censuses and Surveys Classification of Surgical Operations and Procedures, version four (OPCS-4). This classification contains hierarchical codes for interventions and procedures undertaken by the NHS. We removed codes beginning with “F” within ICD-10, which relate to psychiatric or neurological disorders, or both, as this was our outcome of interest. To maintain the hierarchical structure of diagnoses and interventions, we assigned different features to each class level (e.g. ‘G1’,’G12’,’G12.1’) and used one-hot encodings with each unique encoding referring to presence versus absence of a particular diagnosis. To maintain a manageable level of sparsity while retaining the largest amount of useful clinical information, only diagnosis and intervention codes with a prevalence within the cohort of 2% or greater were retained. If a diagnosis did not meet this threshold, it was still included via all parent classes that qualify (e.g. G12.1 had a prevalence below the threshold of 0.4%, but its parent class G12 had a frequency of 6%, so was retained).

### Risk Factors

To improve understanding of mental health risk factors, we utilised a framework previously developed by the team through a rapid review and Delphi process (manuscript in draft). The framework contains 287 risk factors, grouped into seven domains: social and environmental, behavioural, education and employment, biomarkers, physical health, psychological and mental health, and patterns of service use (Supplemental Table 2). An eighth domain combines the risk factors from these domains that are particularly relevant for underserved populations. A mapping exercise between the Delphi risk factor framework to SAIL data established that 101 of the 287 (35.19%) were measurable. Of these, 41 met the missing values criteria of having data for at least 20% of the cohort and were included in the final model. Some of these risk factors (e.g. ethnicity) have multiple categories, thus there are more categorical features in the model than original risk factors. The final risk factors included correspond to six continuous features and 69 categorical features in the model. Exploration of comorbid diagnoses and chronic medical conditions from the Patient Episode Database for Wales (PEDW) as risk factors yielded 2,643 unique diagnostic codes and 1,185 unique intervention codes. 83.04% of children had at least one diagnosis listed and 55.08% had at least one intervention listed. 61 unique diagnoses and 23 unique interventions met the 2% prevalence cut-off and were included within the model as features. Together, these provided 159 features that were used for modelling.

Risk factors with values at multiple time points were converted into binary variables indicating whether the individual had ever been exposed to the risk factor. For children with a mental health diagnosis, risk factor data were only included if it occurred temporally prior to the first positive recorded instance of a mental health problem. For children without a mental health diagnosis, all information was included for prediction up to the final date that they had social care data. Given this is a real-world clinical dataset, there is substantial missing data. If there were missing data regarding a risk factor, individuals were categorised as “Unknown” for that risk factor, and this was included as a feature for the models. This approach was chosen because it provides full flexibility to the models by allowing them to weigh the importance of missing data. Advantages and limitations of this approach are explored in the discussion.

To reduce multicollinearity and subsequently improve interpretability, categorical risk factors were represented as one-hot encodings. To support interpretation of regression modelling, the reference category for each risk factor was coded as the category assumed to have the lowest association with diagnosis of a mental health problem (e.g. reference category for parents’ smoking status would be “non-smoker”). Continuous variables were standardised using sample means and standard deviations with absolute cut-offs applied at ±4 standard deviations from the mean to remove errors and extreme anomalies. If a value for a continuous variable was missing, it was set to the mean of that variable. If a continuous variable had some missing data, an additional binary variable was created to indicate missing data. Some risk factors related to an individual’s early life and child development (e.g. birth weight), while some related to demographic data (e.g. age), and others related to difficult childhood experiences (e.g. abuse). Taken together, these variables constitute risk factors relevant at different stages of an individual’s life course. Including ethnicity in a predictive model that supports decision-making regarding care access has potential equity ramifications. However, given our exploratory focus, we retained ethnicity data to gain insight into how to create equitable classifiers.

### Modelling Decisions

Many ML methods applied to clinical datasets have shown success by utilising Recurrent Neural Network (RNN) model structures to model time-series data (18–20). However, the majority of our datasets are derived from annual censuses that are not sufficiently granular to merit a time-series analysis. Prior research demonstrated the labels of a psychiatry diagnosis are insufficient for modelling since psychiatric diseases are often heterogeneous, multifactorial, and highly comorbid (12,13). Further, transdiagnostic interventions relating to prevention and treatment of childhood mental health problems demonstrate efficacy regardless of the underlying pathology (21). Thus, we framed the prediction task as a binary classification problem (i.e. the presence or absence of a mental health problem).

Within the cohort, a minority of the children have a mental health diagnosis, so a model could theoretically achieve high accuracy by classifying all individuals as healthy. Since such a model would not have clinical utility, loss functions were adjusted to apply greater emphasis (weight) to the correct classification of children with a mental health diagnosis. The standard formulae for weighting to obtain balanced class performances are shown in Equation 1 for those with a mental health problem (MH^+^) and Equation 2 for those without (MH^-^).

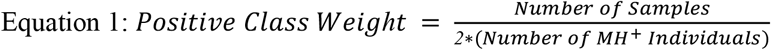

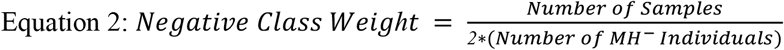

Due to its interpretability, logistic regression was used as the baseline model. Other standard models implemented included Support Vector Machines (SVM), Random Forest models, Multilayer Perceptrons (MLP), and Gradient Boosting models. These additional models are more complex than logistic regression and have associated model “hyperparameters” (e.g. size of the model) whose values are fixed before the model is trained. To find the optimal hyperparameter values, we performed a grid search using the validation set. The MLP model was created using PyTorch (22), while the remaining models were created using Scikit-Learn (23). No feature selection was performed for the models. The best-performing models of each class were then evaluated on the test set. The hyperparameter search space and values of the optimised hyperparameters are shown in Supplementary Table 3.

### Performance Metrics

Due to our unbalanced dataset, we use area under the receiver operating characteristic curve (AUROC) as the primary evaluation metric. AUROC can be interpreted as the probability that a classifier will rank a randomly chosen positive instance higher than that of a randomly chosen negative instance (24). We also report area under the precision–recall curve (AUPRC) as a supplementary evaluation metric. 95% confidence intervals (CI) for AUROC and AUPRC were calculated by using bootstrapping to resample the test set 500 times. This is a required step to obtain confidence estimates since the performance of deterministic models such as logistic regression does not vary with different training iterations.

### Fairness Metrics

We utilise common fairness metrics (equalised odds and predictive parity) to gain insights into model performance for populations that differ with regards to two salient characteristics: biological sex and ethnicity. Equalised odds parity is satisfied when the True Positive Rate (TPR), also known as sensitivity, and the True Negative Rate (TNR), also known as specificity, are equivalent for the groups of interest. Predictive parity, in contrast, is satisfied when the Positive Predictive Value (PPV) and Negative Predictive Value (NPV) are equivalent for the groups of interest (25).

### Ethics

Our application to obtain access to SAIL was reviewed and approved by the internal and external Information Governance Review Panel (IGRP). Since all datasets are anonymised and there is statistical disclosure control for outputs (e.g. reported results must include a minimum of five individuals), there is no legal requirement for the obtainment of individual consent.

## Results

### Cohort Description

The baseline cohort included a sample of 1,113,776 children, of which 46,744 had social care contact. Individuals were excluded if they were under 10 years old (17,992; 38.49%), could not be linked to the other datasets (1,753; 3.75%), or data regarding their mental health status was not available (149; 0.32%). This reduced the final cohort size to 26,820 individuals (57.38%). There were 18,774 individuals in the training dataset, 4,023 in the validation set, and 4,023 in the test set. Demographic information is shown in Table 1.

**Table 1:** Cohort Demographics.

| Variable | Total Dataset | MH<sup>+</sup> Children | MH<sup>-</sup> Children | $\chi^2$ | P-value |
| --- | --- | --- | --- | --- | --- |
| **Sample Size** | 26,820 (100%) | 5,303 (19.77%) | 21,517 (80.23%) |  |  |
| **Age** |  |  |  | 300 | <.00001 |
| 10-12 | 8,845 (32.98%) | 1,374 (25.91%) | 7,471 (34.72%) | 150 | <.00001 |
| 13-15 | 9,801 (36.54%) | 2,441 (46.03%) | 7,360 (34.21%) | 257 | <.00001 |
| 16-18 | 7,868 (29.34%) | 1,466 (27.64%) | 6,402 (29.75%) | 9.12 | <.01 |
| 19-21 | 306 (1.14%) | 22 (0.41%) | 284 (1.32%) | 30.9 | <.00001 |
| **Biological Sex** |  |  |  | 34.5 | <.00001 |
| Female | 14,283 (53.26%) | 2,633 (49.65%) | 11,650 (54.14%) |  |  |
| Male | 12,537 (46.74%) | 2,670 (50.35%) | 9,867 (45.86%) |  |  |
| **Ethnicity** |  |  |  | 39.0 | <.00001 |
| Asian | 485 (1.81%) | 56 (1.06%) | 429 (1.99%) | 21.1 | <.00001 |
| Black | 314 (1.17%) | 45 (0.85%) | 269 (1.25%) | 5.93 | <.05 |
| Mixed | 696 (2.60%) | 139 (2.62%) | 557 (2.59%) | 0.018 | .894 |
| White | 24,010 (89.52%) | 4,840 (91.27%) | 19,170 (89.09%) | 21.5 | <.00001 |
| Other | 263 (0.98%) | 36 (0.68%) | 227 (1.05%) | 6.20 | <.05 |
| Not Obtained | 886 (3.30%) | 163 (3.07%) | 723 (3.36%) | 1.09 | .296 |
| Refused | 166 (0.62%) | 24 (0.45%) | 142 (0.66%) | 2.97 | .085 |
| **Free School Meal Status** |  |  |  | 8.59 | <.05 |
| Eligible | 20,253 (75.51%) | 3,922 (73.96%) | 16,331 (75.90%) | 7.83 | <.01 |
| Not Eligible | 6,460 (24.09%) | 1,360 (25.65%) | 5,100 (23.70%) | 8.79 | <.01 |
| Unknown | 107 (0.40%) | 21 (0.40%) | 86 (0.40%) | 0.0009 | .977 |

The mean age among children who experienced a mental health problem was 14.5 years (SD 2.15). There was a higher prevalence of mental health problems in males (21.30%) compared to females (18.43%). Given the class imbalance, the weight given by Equation 1 was 2.53 and the weight given by Equation 2 was 0.62, corresponding to upweighting the class of individuals with a mental health problem by 4.06. The most common ethnicity within the dataset was white (89.52%). The demographics of this dataset are similar to the overall demographics of Wales recorded in the 2011 Census (26).

### Model Interpretability

The 20 highest statistically significant coefficients for the logistic regression model are shown in Fig. 1, demonstrating the variables most closely associated with adverse mental health outcomes. 95% confidence intervals for the logistic regression model were calculated by bootstrapping to resample the training set 500 times.

**Fig. 1:**
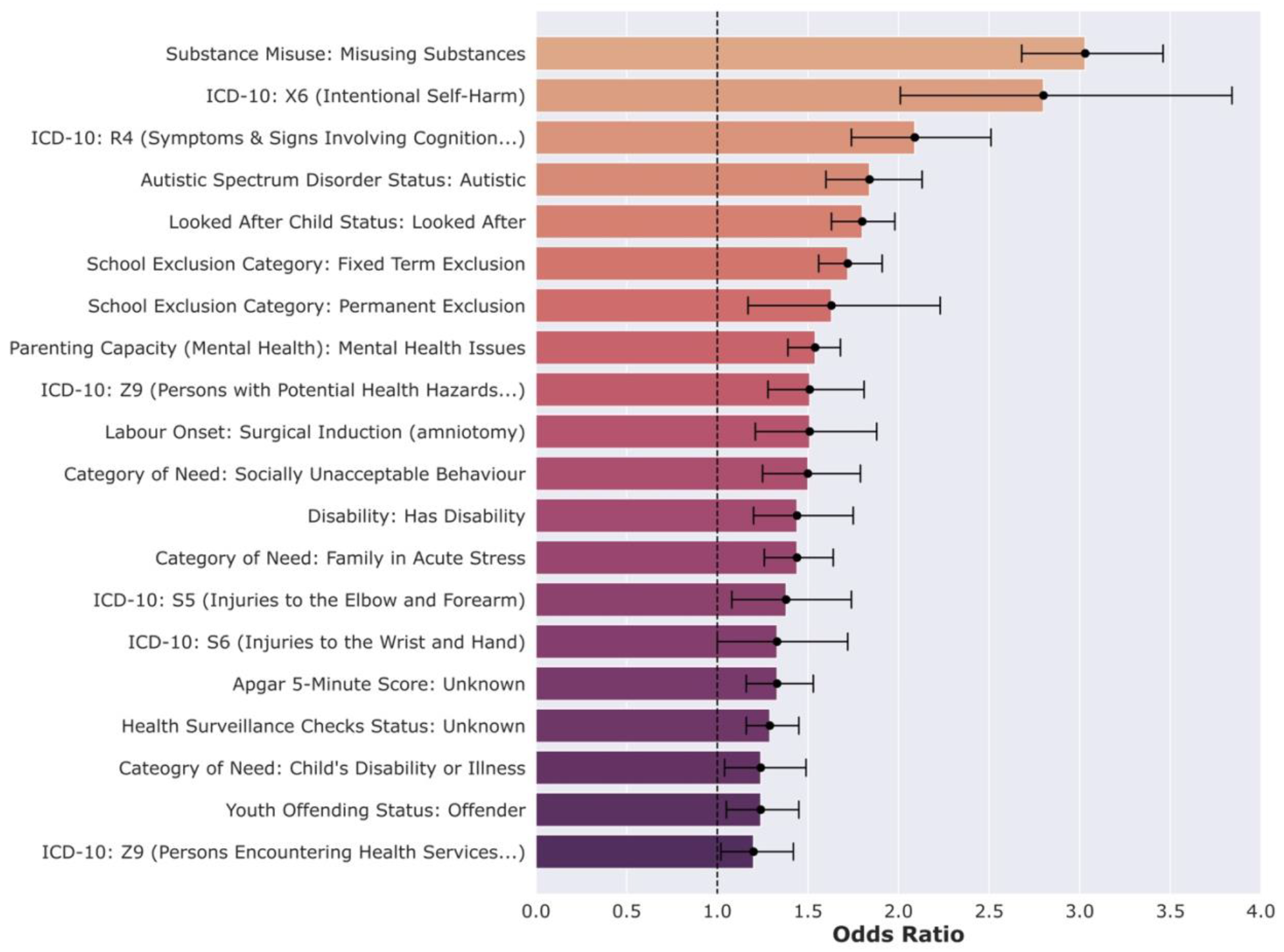
Odds Ratios for Interpretable Logistic Regression Model.

The five risk factors with the largest odds ratios were: ‘Substance Misuse: Misusing Substances,’ ‘ICD-10 Code: X6 (Intentional Self-Harm),’ ‘ICD-10 Code: R4 (Symptoms and Signs Involving Cognition, Perception, Emotional State and Behaviour),’ ‘Autistic Spectrum Disorder Status: Autistic,’ and ‘Looked After Child Status: Looked After.’ Odds ratios above one indicate that the feature is associated with a greater risk of being diagnosed with a mental health problem. The complete list of 159 feature coefficients for the logistic regression model is shown in Supplemental Table 4.

### Model Performance

The performance of the models on the validation and test datasets is shown in Table 2.

**Table 2:** Model Performance on Validation (Val) and Test Datasets with 95% Confidence Interval.

| Model | AUROC Val | AUROC Test | AUPRC Val | AUPRC Test |
| --- | --- | --- | --- | --- |
| Logistic Regression | 0.715 (0.694-0.734) | 0.703 (0.682-0.724) | 0.405 (0.369-0.439) | 0.395 (0.363-0.429) |
| Support Vector Machine (SVM) | 0.750 (0.732-0.770) | 0.732 (0.712-0.751) | 0.449 (0.413-0.483) | 0.443 (0.409-0.477) |
| Random Forest | 0.730 (0.711-0.750) | 0.705 (0.683-0.724) | 0.409 (0.374-0.444) | 0.405 (0.370-0.437) |
| Gradient Boosting Classifier | 0.749 (0.730-0.768) | 0.685 (0.664-0.706) | 0.455 (0.418-0.489) | 0.369 (0.334-0.406) |
| Multilayer Perceptron (MLP) | 0.743 (0.724-0.762) | 0.724 (0.703-0.743) | 0.436 (0.402-0.472) | 0.432 (0.398-0.465) |

The best-performing model on the test dataset is the SVM model which achieved an AUROC of 0.732 and an AUPRC of 0.443, while the next best-performing model is the MLP model with an AUROC of 0.724 and an AUPRC of 0.432. Models tended to perform better on the validation set than they performed on the test dataset, which is expected for ML models tuned on the validation set. There is a high degree of concordance between the AUROC and AUPRC. The Gradient Boosting Classifier did not generalise well to the test dataset, with the AUROC dropping from 0.749 on the validation dataset to 0.685 on the test dataset.

### Algorithmic Fairness

For the best-performing model (SVM model), assessments of algorithmic fairness are displayed in Fig. 2. These results in tabular form can be seen in Supplemental Table 5. For all four fairness metrics, values closer to one signify better performance, while values closer to zero signify worse performance.

**Fig. 2:**
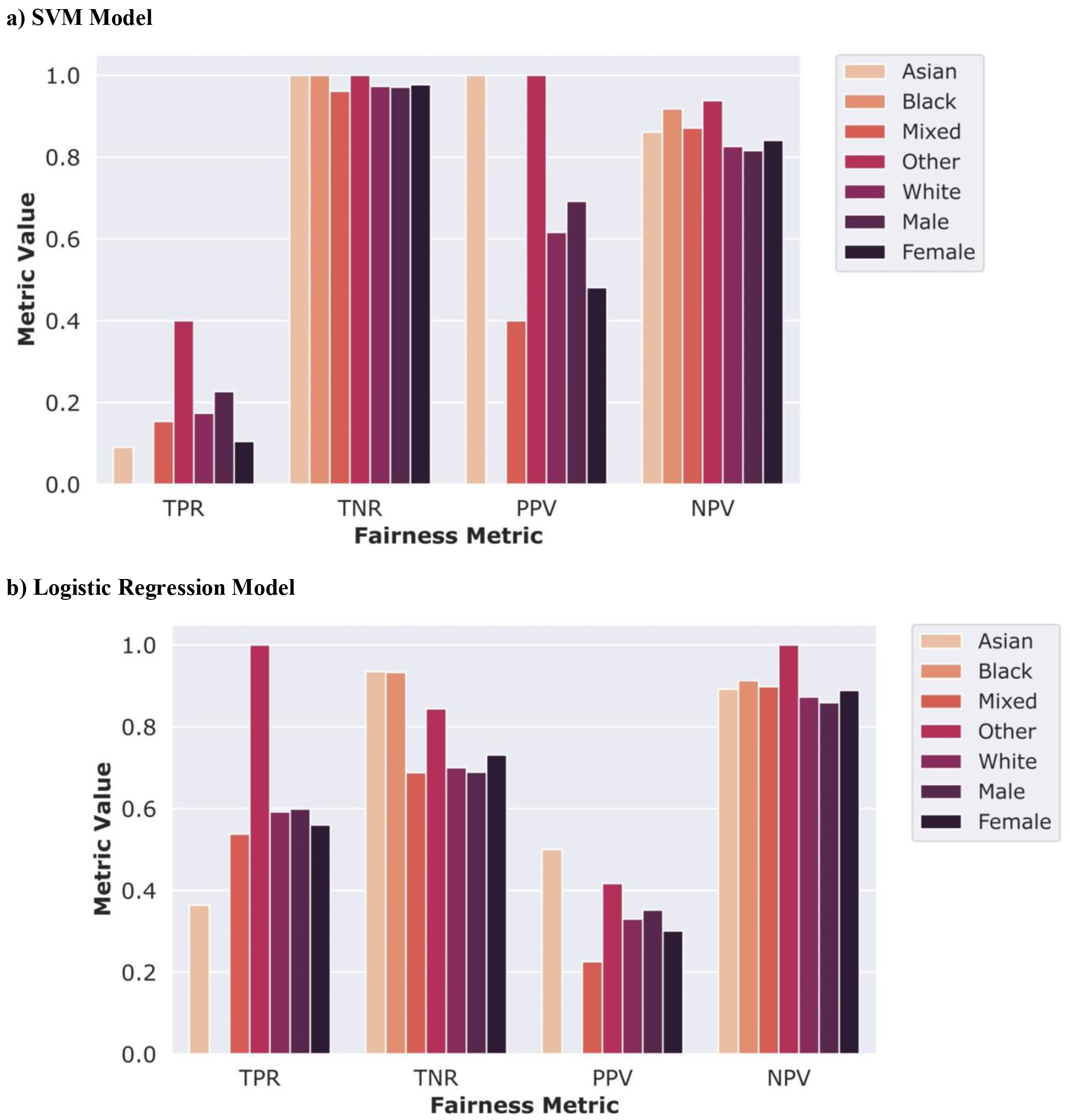
Assessment of Algorithmic fairness.

The SVM model has a high TNR and low TPR for all ethnic and sex groups, indicating that it is better at correctly predicting individuals who do not have a mental health problem and has more difficulty correctly predicting individuals with a mental health problem. In contrast, the logistic regression model more effectively balances the two classes with similar results for TPR and TNR. The SVM model exhibits similar TNR and NPV performance for all ethnicities and biological sex but shows more variance in TPR and PPV. In both models, there is possible sex bias with a higher TPR and PPV for males and a higher TNR and NPV for females.

## Discussion

### Main Findings

Within the cohort, there is a higher prevalence of mental health problems compared to the general population of children in the UK. This likely reflects that children in social care experience more ACEs, known to be associated with a range of mental health issues (27). Many of the risk factors most correlated with a mental health diagnosis are already well-established including substance misuse, having a disability, and an autism comorbidity. Other mental health risk factors identified are less-established and deserve additional examination, such as surgical induction of labour in the mother during birth of the child and injuries to the arm (ICD-10 S5 and S6).

The most prominent risk factors were an amalgamation of personal risk factors (e.g. having a disability) and risk factors related to family (e.g. mental health of parents). The presence of missing data for two features (Apgar score, and health surveillance checks) was significantly associated with a mental health diagnosis. Missing data is difficult to interpret because observed changes could be attributable to individual-level factors or factors relating to data collection. For mental health risk factors, it is possible that missing data for certain risk factors such as child health surveillance checks could be tied to patterns of service use (i.e. indicative of children’s routine health checks being missed). Further, many of the missing data features had odds ratios with wide confidence intervals, likely reflecting the small sample sizes of missing data classes and the heterogeneity of individuals with missing data. Further analyses could explore additional missing data methods and their relative impacts on the trade-off between interpretability and model performance.

The models achieved good performance on the held-out test dataset, meriting consideration for future use. These results demonstrate that integrating disparate datasets can create powerful prediction models for children in social care settings. However, the models developed require refinement before clinical implementation, such as assessing whether models may exhibit better performance for specific mental health conditions. Model performance did not directly scale with model complexity. For instance, although the logistic regression model was outperformed by most other models, it performed better than the Gradient Boosting Classifier, a more complex model. Additionally, the MLP model (the most complex model assessed) performed worse than the less complex SVM model.

Preliminary assessment of algorithmic fairness illustrated that model performance trends between ethnicities were difficult to disentangle due to small sample sizes for many ethnicities within the test set. For instance, there were only 49 black children in the test dataset, of which four had a mental health diagnosis. Nonetheless, it is notable that no black children with a mental health diagnosis were correctly identified by the model. For biological sex, both models more often correctly identified mental health problems (i.e. true positives) in males than females, while more often identifying lack of mental health problems (i.e. true negatives) in females than males. This trend can be partially explained by the higher prevalence of mental health problems in males (21.30%) than females (18.43%). However, this does not fully account for this discrepancy, and additional factors such as model bias are likely involved. Further evaluation of model fairness is necessary to ensure these models do not exacerbate healthcare disparities.

### Limitations

There are limitations relevant to both cohort creation and model development. The lack of timestamp granularity in SAIL prohibited us from modelling the data using time-series approaches. Further, the SAIL metadata quality was sometimes unclear, forcing us to omit otherwise useful indicators such as urbanicity. Moreover, by focusing solely on children with social care encounters in Wales, generalisability to other populations is diminished. However, this work may be useful for the Welsh population and can still serve as an effective guide when developing more generalisable models.

Importantly, some children with mental health problems either do not seek support services or are unable to access them (28), and consequently cannot be identified with this paper’s methods. Further exacerbating biases, outcome labels are likely skewed towards mental health diagnoses for individuals with more severe mental health problems. Our cohort also lacked parental information, which may limit access to important risk factors. For example, in a similar study, 72.3% of ACEs were found only in maternal records (29). SAIL affords the ability to link in parents’ data with an application amendment, so this remains a potential avenue of future research.

One major modelling limitation is not performing an external validation of model performance. Although we applied proper internal validation safeguards on a large dataset, external validations remain the gold standard. Finally, the standardised weighting factor was insufficient to ensure that all models effectively balance sensitivity and specificity. This limits the clinical effectiveness of the models, and future work should explore varying the weighting factor.

### Implications and Future Work

This work comprises one portion of an overarching project to create CADRE (Child and Adolescent Data REsource), a database containing administrative data relating to health, social care, and education for young people aged 0 to 17 years. The aim is for CADRE to be usable for real-time clinical decision support, with a de-identified version available to approved researchers. CADRE will expand on SAIL’s offering by building a network of Trusted Research Environments across Cambridgeshire, Peterborough, Birmingham, and Essex that will contain genetic data on children and house unstructured data such as clinical notes. The models prototyped within this work will be refined and externally validated in the CADRE database.

There is scant prior work using predictive modelling to identify mental health problems in children, with a recent systematic review (30) finding only two articles meeting these criteria (9,31). Although difficult to directly compare results, model performance here is on par with previous analyses. This work builds upon these previous analyses by assessing a substantially larger cohort of 26,820 children (prior two studies looked at 7,638 and 60 children). In this work, we also identified mental health risk factors that healthcare professionals should consider when caring for children, especially those with social care contact. This analysis also details ML techniques including assessments of algorithmic fairness useful for future related work.

## Data Availability

The raw data used for this study is housed by the SAIL Databank. This databank is not available publicly, but researchers can access the data following approval by the SAIL IGRP. Information regarding this application process can be found at https://saildatabank.com/application-process/.

## Acknowledgements

We would like to thank the Secure Anonymised Information Linkage (SAIL) Databank, and in particular all the individuals whose data makes this data safe haven possible. We would like to thank the Adolescent Mental Health Data Platform (ADP) including Hannah Evans and Marcos Del Pozo Banos for providing guidance. The ADP is funded by MQ Mental Health Research Charity (Grant Reference MQBF/3 ADP). ADP and the author(s) would like to acknowledge the data providers who supplied the datasets enabling this research study. The views expressed are entirely those of the authors and should not be assumed to be the same as those of ADP or MQ Mental Health Research Charity. We would also like to thank Dr. Angela Wood, and Prof. Zoe Kourtzi for providing supervision and guidance for the machine learning aspects of this project. Finally, we would like to thank our funders, without whom this work would not have been possible.

## Author Contributions

R.C., K.P., Y.F., A.J., and E.M curated the database. R.C., E.R, K.P., R.S., A.M., P.L., and E.M. were involved in the analysis. All authors made substantial contributions to the design of the work, helped interpret the data, revised the work regarding important intellectual content, and provided final approval of the submitted manuscript.

