## Supplemental Data for "Machine Learning for Prediction of Childhood Mental Health Problems in Social Care"

### Appendix

**Table 1: SAIL Datasets**

| Group Name | Individual Data Sources | Included in Final Analysis |
|---|---|---|
| Demographics | Annual District Birth Extract (ADBE) | N |
| | Annual District Death Extract (ADDE) | N |
| | Welsh Demographic Service Dataset (WDSD) | Y |
| Education | Pre16 Education Attainment (EDUW) | Y |
| GP | GP Primary Care – Audit (WLGP) | N |
| Healthcare | Critical Care Dataset (CCDS) | N |
| | Emergency Department Dataset (EDDS) | N |
| | Maternity Indicators Dataset (MIDS) | Y |
| | National Community Child Health (NCCH) | Y |
| | NHS Hospital Outpatients (OPDW) | N |
| | NHS 111 Call data (NHSO) | N |
| | Outpatient Referrals from Primary Care (OPRD) | N |
| | Patient Episode Database for Wales (PEDW) | Y |
| | Substance Misuse Dataset (SMDS) | N |
| | Wales Results Reporting Service (WRRS) | N |
| Social Care | Child In Need Wales (CINW) | Y |
| | Children Receiving Care and Support (CRCS) | Y |
| | Looked After Children Wales (LACW) | N |

**Table 2: Complete Risk Factors from Delphi**

| Risk Factor | Domain | Level |
|---|---|---|
| Air pollution | Social and Environmental | Society |
| Area deprivation (area code) | Social and Environmental | Society |
| Ethnic minority in low ethnic density area | Social and Environmental | Society |
| English as an additional language | Social and Environmental | Society |
| Gangland crime | Social and Environmental | Society |
| Remoteness of living | Social and Environmental | Society |
| Urbanicity | Social and Environmental | Society |
| War/conflict | Social and Environmental | Society |
| Broken or complex family structure | Social and Environmental | Family/caregiver |
| Death of close relative (excluding primary caregiver(s)) | Social and Environmental | Family/caregiver |
| Death of primary caregiver(s) | Social and Environmental | Family/caregiver |
| Domestic violence | Social and Environmental | Family/caregiver |
| Family conflict or discord | Social and Environmental | Family/caregiver |
| Family financial problems | Social and Environmental | Family/caregiver |
| Famine/food poverty | Social and Environmental | Family/caregiver |
| High socioeconomic status family | Social and Environmental | Family/caregiver |
| Household alcohol abuse | Social and Environmental | Family/caregiver |
| Household criminality | Social and Environmental | Family/caregiver |

| | | |
|---|---|---|
| Household drug abuse | Social and Environmental | Family/caregiver |
| Household mental illness | Social and Environmental | Family/caregiver |
| Household overcrowding | Social and Environmental | Family/caregiver |
| Low social support for family | Social and Environmental | Family/caregiver |
| Low socioeconomic status family | Social and Environmental | Family/caregiver |
| Multi-generational families within the same home | Social and Environmental | Family/caregiver |
| Number of living children in the family | Social and Environmental | Family/caregiver |
| Poor primary caregiver-caregiver relationship | Social and Environmental | Family/caregiver |
| Poor relationship between primary caregivers | Social and Environmental | Family/caregiver |
| Primary caregiver(s) marital status | Social and Environmental | Family/caregiver |
| Primary caregiver(s) parenting styles (strict/rigid/conventional) | Social and Environmental | Family/caregiver |
| Primary caregiver(s) unemployment | Social and Environmental | Family/caregiver |
| Single parent/caregiver family | Social and Environmental | Family/caregiver |
| Smoking status of primary caregiver(s) | Social and Environmental | Family/caregiver |
| Smoking status of primary caregiver(s) | Social and Environmental | Family/caregiver |
| Being a second generation immigrant | Social and Environmental | Individual |
| Being a young carer | Social and Environmental | Individual |
| Child younger than classmates | Social and Environmental | Individual |
| Chronic psychological stress | Social and Environmental | Individual |
| Cumulative traumatic events | Social and Environmental | Individual |

| | | |
|---|---|---|
| Death of a close friend | Social and Environmental | Individual |
| Emotional neglect | Social and Environmental | Individual |
| Emotional, psychological or verbal abuse | Social and Environmental | Individual |
| Ethnicity | Social and Environmental | Individual |
| Experience of past traumatic event | Social and Environmental | Individual |
| Frequent fear of family member | Social and Environmental | Individual |
| Experience of racism and discrimination | Social and Environmental | Individual |
| Injury during a traumatic event | Social and Environmental | Individual |
| Maternal alcohol use during pregnancy | Social and Environmental | Individual |
| Maternal smoking during pregnancy | Social and Environmental | Individual |
| Maternal substance abuse during pregnancy | Social and Environmental | Individual |
| Maternal use of psychotropics during pregnancy | Social and Environmental | Individual |
| Month of birth | Social and Environmental | Individual |
| Not breast fed | Social and Environmental | Individual |
| Perceived pressure to be thin | Social and Environmental | Individual |
| Physical abuse | Social and Environmental | Individual |
| Physical neglect | Social and Environmental | Individual |
| Poor online peer relationships | Social and Environmental | Individual |
| Poor peer relationships | Social and Environmental | Individual |
| Poor primary caregiver-child relationship | Social and Environmental | Individual |
| Separation from family (e.g. out-of-home care) | Social and Environmental | Individual |

| | | |
|---|---|---|
| Sexual abuse | Social and Environmental | Individual |
| Torture | Social and Environmental | Individual |
| Trapped during earthquake | Social and Environmental | Individual |
| Trauma severity | Social and Environmental | Individual |
| Victim of bullying | Social and Environmental | Individual |
| Victim or witness of violent crime | Social and Environmental | Individual |
| Witness of community violence | Social and Environmental | Individual |
| Witnessing injury/death during a traumatic event | Social and Environmental | Individual |
| Physical inactivity in family/primary care-givers | Behavioural | Family/caregiver |
| Unhealthy diet in family/primary care-givers | Behavioural | Family/caregiver |
| Diet low in plant-matter | Behavioural | Individual |
| Diet low in plant-matter diversity | Behavioural | Individual |
| Excessive physical activity | Behavioural | Individual |
| Excessive sports participation | Behavioural | Individual |
| Foods high in trans fats in diet | Behavioural | Individual |
| Frequent social media use | Behavioural | Individual |
| Heavy alcohol use | Behavioural | Individual |
| High ratio of omega-6 to omega-3 fatty acids in diet | Behavioural | Individual |
| Higher behavioural inhibition | Behavioural | Individual |
| Lower behavioural inhibition | Behavioural | Individual |

| | | |
|---|---|---|
| Low levels of omega-3 polyunsaturated fatty acids in diet | Behavioural | Individual |
| Low sports participation | Behavioural | Individual |
| Nitrous oxide use | Behavioural | Individual |
| Non-prescription drug use | Behavioural | Individual |
| Physical inactivity | Behavioural | Individual |
| Poor sleep patterns | Behavioural | Individual |
| Prescription drug abuse | Behavioural | Individual |
| Sexual risk-taking | Behavioural | Individual |
| Smoking | Behavioural | Individual |
| High levels of ultra-processed foods in diet | Behavioural | Individual |
| Unhealthy diet | Behavioural | Individual |
| Increased pressure/stress for teachers (e.g. insufficient pay/resources and poor leadership) | Education/Employment | Society |
| Poor school climate (e.g. Ofsted weightings) | Education/Employment | Society |
| Poor quality of a lack of social-emotional learning programmes in pre-school | Education/Employment | Society |
| Quality of school climate (e.g. relating to school connectedness, feelings of safety in school, perception of school, adult-student relationships, morale) | Education/Employment | Society |
| School composition (e.g. size/headcount, gender proportions, ethnicity proportions) | Education/Employment | Society |

| | | |
|---|---|---|
| School-level deprivation (e.g. proportion eligible for free school meals) | Education/Employment | Society |
| Language impairment in primary caregivers | Education/Employment | Family/caregiver |
| Low education level of caregiver(s) (other than primary caregiver(s)) | Education/Employment | Family/caregiver |
| Low education level of primary caregiver(s) | Education/Employment | Family/caregiver |
| Child not read to daily by age 1 | Education/Employment | Individual |
| Disruptive behaviours in school (e.g. defiance and non-compliance) | Education/Employment | Individual |
| Having an Education and Health Care Plan (EHCP) | Education/Employment | Individual |
| Lack of imitation games with caregivers by age 1 | Education/Employment | Individual |
| Out-of-school discipline (e.g. suspension and expulsion) | Education/Employment | Individual |
| Participation in the Free-Lunch Program | Education/Employment | Individual |
| Poor educational attainment | Education/Employment | Individual |
| Poor peer relationships | Education/Employment | Individual |
| Poor school attendance | Education/Employment | Individual |
| Poor teacher-pupil relationship | Education/Employment | Individual |
| School exclusions | Education/Employment | Individual |
| Special Educational Needs (SEN) | Education/Employment | Individual |
| Unemployment of the individual | Education/Employment | Individual |
| Age of parents at time of giving birth | Biomarkers | Individual |
| Birth length | Biomarkers | Individual |

| | | |
|---|---|---|
| Birth weight | Biomarkers | Individual |
| CD14 levels | Biomarkers | Individual |
| Changes in amygdala (e.g. reduced volume) | Biomarkers | Individual |
| Changes in prefrontal cortex (e.g. reduced total volume; low myelination; low white matter; increased middle inferior and ventral regions and superior/dorsal regions) | Biomarkers | Individual |
| Chronic inflammation | Biomarkers | Individual |
| Decreased neuronal branching | Biomarkers | Individual |
| Epigenetic marker - chromatin regulation | Biomarkers | Individual |
| Epigenetic marker - DNA methylation | Biomarkers | Individual |
| Epigenetic marker - non-coding ribonucleic acids | Biomarkers | Individual |
| Genetic markers | Biomarkers | Individual |
| Gut microbiome (reduced diversity and reduced populations of certain bacteria) | Biomarkers | Individual |
| High intelligence quotient (IQ) | Biomarkers | Individual |
| High levels of cytokine proteins (e.g. interleukin-1 [IL-1]; interleukin-6 [IL-6]) | Biomarkers | Individual |
| High levels of indoleamine-2,3-dioxygenase (IDO) | Biomarkers | Individual |
| High levels of tryptophan-2,3-dioxygenase (TDO) | Biomarkers | Individual |
| High peripheral manganese | Biomarkers | Individual |
| High permeability of blood-brain barrier | Biomarkers | Individual |

| | | |
|---|---|---|
| HPA axis and glucocorticoid receptor resistance | Biomarkers | Individual |
| Increased levels of chemokines | Biomarkers | Individual |
| Increased levels of eicosanoids | Biomarkers | Individual |
| Increased neurodegeneration | Biomarkers | Individual |
| Increased quinolinic acid (QUIN) | Biomarkers | Individual |
| Kynurenine/tryptophan ratio | Biomarkers | Individual |
| Low levels of dopamine | Biomarkers | Individual |
| Low or high levels of GDNF (i.e. neurotrophic factor) | Biomarkers | Individual |
| Low or high levels of Tumour Necrosis Factor alpha (TNF-α) | Biomarkers | Individual |
| Low intelligence quotient (IQ) | Biomarkers | Individual |
| Low levels of B vitamins | Biomarkers | Individual |
| Low levels of folate | Biomarkers | Individual |
| Low levels of melatonin | Biomarkers | Individual |
| Low levels of nutrients | Biomarkers | Individual |
| Low levels of serotonin | Biomarkers | Individual |
| Low levels of vitamin D | Biomarkers | Individual |
| Low serum ferritin | Biomarkers | Individual |
| Neuronal atrophy | Biomarkers | Individual |
| Neurotoxicity | Biomarkers | Individual |
| Neutrophil–lymphocyte ratio (NLR) | Biomarkers | Individual |

| | | |
|---|---|---|
| Nutritional deficits | Biomarkers | Individual |
| Raised C-reactive protein (CRP) | Biomarkers | Individual |
| Raised levels of glucocorticoids/cortisol | Biomarkers | Individual |
| Reduced grey matter in hippocampus | Biomarkers | Individual |
| Reduced kynurenic acid (KYNA) | Biomarkers | Individual |
| Reduced levels of BDNF (i.e. neurotrophic factor) | Biomarkers | Individual |
| Reduced neurogenesis | Biomarkers | Individual |
| Reduced neuroplasticity | Biomarkers | Individual |
| Sex (biological) | Biomarkers | Individual |
| Family history of autoimmune disease | Physical Health | Family/caregiver |
| Family history of psoriasis | Physical Health | Family/caregiver |
| Family history of rheumatoid arthritis | Physical Health | Family/caregiver |
| Family history of Type 1 diabetes | Physical Health | Family/caregiver |
| Maternal self-rated health (prior to pregnancy) | Physical Health | Family/caregiver |
| Primary caregiver(s) chronic (long lasting) illness | Physical Health | Family/caregiver |
| Severe illness in family | Physical Health | Family/caregiver |
| 5-min Apgar score <7 | Physical Health | Individual |
| Allergies (e.g. non-lgE-mediated food allergies, pollen allergies) | Physical Health | Individual |
| Anaemia | Physical Health | Individual |
| Autoimmune disorders (e.g. rheumatoid arthritis) | Physical Health | Individual |

| | | |
|---|---|---|
| Asthma | Physical Health | Individual |
| Body mass Index (BMI) | Physical Health | Individual |
| Eczema | Physical Health | Individual |
| Chronic (long lasting) gastric ill-health (e.g. inflammatory bowel disease) | Physical Health | Individual |
| Chronic (long lasting) infection (e.g. Lyme disease, periodontal disease) | Physical Health | Individual |
| Chronic (long lasting) reflux or indigestion | Physical Health | Individual |
| Congenital malformations | Physical Health | Individual |
| Dental caries (tooth decay) | Physical Health | Individual |
| Dental erosion | Physical Health | Individual |
| Diabetes | Physical Health | Individual |
| Disease history | Physical Health | Individual |
| Global developmental delay | Physical Health | Individual |
| Hypoxia (at birth) | Physical Health | Individual |
| Inflammatory diseases (e.g. Lyme disease) | Physical Health | Individual |
| Irritable Bowel Syndrome (IBS) | Physical Health | Individual |
| Lack of response to treatment for a physical health condition | Physical Health | Individual |
| Learning disability | Physical Health | Individual |
| Long term antibiotic use | Physical Health | Individual |
| Low diversity & composition of gut microbiota | Physical Health | Individual |

| | | |
|---|---|---|
| Low serum vitamin D | Physical Health | Individual |
| Maternal hypertension during pregnancy | Physical Health | Individual |
| Maternal infection requiring hospitalisation during pregnancy | Physical Health | Individual |
| Maternal obesity during pregnancy | Physical Health | Individual |
| Maternal acetaminophen (paracetamol) use during pregnancy | Physical Health | Individual |
| Maternal auto-immune disease during pregnancy | Physical Health | Individual |
| Maternal diabetes during pregnancy | Physical Health | Individual |
| Maternal obesity/overweight during pregnancy | Physical Health | Individual |
| Obesity/overweight | Physical Health | Individual |
| Obstetric complications (other) | Physical Health | Individual |
| Perinatal infections (e.g. cytomegalovirus) | Physical Health | Individual |
| Polyhydramnios | Physical Health | Individual |
| Pre-eclampsia | Physical Health | Individual |
| Premature birth | Physical Health | Individual |
| Prolonged duration of a physical health condition | Physical Health | Individual |
| Repeated infections | Physical Health | Individual |
| Ruptured membranes (at birth) | Physical Health | Individual |
| Severe health condition | Physical Health | Individual |
| Sleep disorder | Physical Health | Individual |
| Thyroid disease | Physical Health | Individual |

| | | |
|---|---|---|
| Traumatic brain injury | Physical Health | Individual |
| Negative attitudes to mental health problems within society an individual lives in | Psychological and MH | Society |
| Family history of psychiatric disorders | Psychological and MH | Family/caregiver |
| Family history of severe mental illness (e.g. psychosis) | Psychological and MH | Family/caregiver |
| Primary caregiver(s) mental health problems | Psychological and MH | Family/caregiver |
| Acute stress disorder (as a predictor of further mental health problems) | Psychological and MH | Individual |
| Acute stress symptoms (anxiety, avoidance or depression -as a predictor of further mental health problems) | Psychological and MH | Individual |
| Anxiety (as a predictor of further mental health problems) | Psychological and MH | Individual |
| Attention (as a predictor of further mental health problems) | Psychological and MH | Individual |
| Dissociation during traumatic experience | Psychological and MH | Individual |
| Emotional reactivity | Psychological and MH | Individual |
| Excessive rumination | Psychological and MH | Individual |
| High levels of perceived stress | Psychological and MH | Individual |
| Increased anxiety arousal | Psychological and MH | Individual |
| Increased panic attacks | Psychological and MH | Individual |
| Ineffective coping strategies | Psychological and MH | Individual |
| Lack of psychological resilience | Psychological and MH | Individual |

| | | |
|---|---|---|
| Maternal depression during pregnancy | Psychological and MH | Individual |
| Maternal psychosis (perinatal or pre-natal) | Psychological and MH | Individual |
| Poor concentration | Psychological and MH | Individual |
| Poor problem-solving abilities | Psychological and MH | Individual |
| Poor visuospatial functioning | Psychological and MH | Individual |
| Problems with memory | Psychological and MH | Individual |
| Psychiatric history | Psychological and MH | Individual |
| Primary caregiver(s) non-attendance at baby groups | Patterns of Service Use | Family/caregiver |
| Child protection record | Patterns of Service Use | Individual |
| Lack of attendance to dental visits | Patterns of Service Use | Individual |
| Missed ante-natal visits | Patterns of Service Use | Individual |
| Missed doctor appointments | Patterns of Service Use | Individual |
| Missed postnatal visits | Patterns of Service Use | Individual |
| Missed routine check-up appointments | Patterns of Service Use | Individual |
| Missed vaccination appointments | Patterns of Service Use | Individual |
| Presentation to multiple services (e.g. different hospitals, GPs, social care etc) | Patterns of Service Use | Individual |
| Repeat hospitalisation | Patterns of Service Use | Individual |
| Area deprivation (area code) | Factors Relevant to Under-Served Populations | Society |
| English as an additional language | Factors Relevant to Under-Served Populations | Society |

| | | |
|---|---|---|
| Ethnic minority in low ethnic density area | Factors Relevant to Under-Served Populations | Society |
| Gangland crime | Factors Relevant to Under-Served Populations | Society |
| Increased pressure/stress for teachers (e.g. insufficient pay/resources and poor leadership) | Factors Relevant to Under-Served Populations | Society |
| Poor school climate (e.g. Ofsted weightings) | Factors Relevant to Under-Served Populations | Society |
| Quality of school climate (e.g. relating to school connectedness, feelings of safety in school, perception of school, adult-student relationships, morale) | Factors Relevant to Under-Served Populations | Society |
| School composition (e.g. size/headcount, gender proportions, ethnicity proportions) | Factors Relevant to Under-Served Populations | Society |
| School-level deprivation (e.g. proportion eligible for free school meals) | Factors Relevant to Under-Served Populations | Society |
| War/conflict | Factors Relevant to Under-Served Populations | Society |
| Family financial problems | Factors Relevant to Under-Served Populations | Family/caregiver |
| Family experience of social exclusion, discrimination and harassment associated with ethnicity | Factors Relevant to Under-Served Populations | Family/caregiver |
| Famine/food poverty | Factors Relevant to Under-Served Populations | Family/caregiver |

| | | |
|---|---|---|
| Household overcrowding | Factors Relevant to Under-Served Populations | Family/caregiver |
| Low socioeconomic status | Factors Relevant to Under-Served Populations | Family/caregiver |
| Multi-generational families within the same home | Factors Relevant to Under-Served Populations | Family/caregiver |
| Primary caregiver(s) parenting styles (strict/rigid/conventional) | Factors Relevant to Under-Served Populations | Family/caregiver |
| Primary caregiver(s) unemployment | Factors Relevant to Under-Served Populations | Family/caregiver |
| Being a second generation immigrant | Factors Relevant to Under-Served Populations | Individual |
| Being a young carer | Factors Relevant to Under-Served Populations | Individual |
| English as an additional language | Factors Relevant to Under-Served Populations | Individual |
| Ethnicity | Factors Relevant to Under-Served Populations | Individual |
| Experience of bereavement during a traumatic event | Factors Relevant to Under-Served Populations | Individual |
| Experience of racism and discrimination | Factors Relevant to Under-Served Populations | Individual |
| Participation in the Free-Lunch Program | Factors Relevant to Under-Served Populations | Individual |
| Trapped during earthquake | Factors Relevant to Under-Served Populations | Individual |

| | | |
|---|---|---|
| Unemployment | Factors Relevant to Under-Served Populations | Individual |

**Table 3: Hyperparameter Tuning by Model**

| **Algorithm** | **Tuning Parameters*** |
|---|---|
| Logistic Regression | N/A |
| SVM (RBF Kernel) | Cost (.1, .25, .5, **1**)<br>Gamma (2e-3, **2e-2** ,2e-1, 1) |
| Random Forest | N_estimators (50, **100**, 150)<br>Max Tree Depth (1, 2, 3, **4**)<br>Max Features (4, 8, **12**, 16) |
| Gradient Boosting Classifier | Number Estimators (50, 100, **150**)<br>Max Depth (1, 2, 3, **4**)<br>Max Features (4, 8, **12**, 16) |
| MLP Classifier | Learning Rate (**1e-4**, 1e-3, 1e-2, 1e-1)<br>L2 (1e-5, 1e-4, **1e-3**)<br>Number of Hidden Layers (1, **2**, 4, 8)<br>Number of Nodes Per Layer (10, 20, 30, **40**) |

*****Bolded hyperparameter values reflect the optimal hyperparameters selected during hyperparameter tuning

**Table 4: Adjusted Odds Ratios with 95% Confidence Interval for Interpretable Logistic Regression Model**

| **Risk Factor** | **Odds Ratio** |
|---|---|
| Age: Continuous Value | 0.78 (0.75-0.82) |
| Apgar 1-Minute Score: Continuous Value | 0.93 (0.87-0.99) |
| Apgar 5-Minute Score: Continuous Value | 1.02 (0.96-1.09) |
| Asylum Seeker Status: Asylum Seeker | 0.93 (0.49-1.69) |
| Asylum Seeker Status: Unknown | 1.19 (0.62-2.1) |
| Birth Weight: Continuous Value | 1.02 (0.97-1.08) |
| Breastfeed Status (8 weeks): Not Breastfed | 0.94 (0.81-1.09) |
| Breastfeed Status (8 weeks): Unknown | 1.04 (0.91-1.22) |
| Breastfeed Status (Birth): Not Breastfed | 1.05 (0.93-1.18) |

| | |
|---|---|
| Breastfeed Status (Birth): Not Breastfed | 0.9 (0.79-1.02) |
| Health Surveillance Checks Status: Not Receiving Checks | 0.93 (0.81-1.05) |
| Health Surveillance Checks Status: Unknown | 1.29 (1.16-1.45) |
| Child Protection Register Status: Registered | 0.9 (0.82-1) |
| Diagnosis Code: A0 (Intestinal Infectious Diseases) | 1.05 (0.92-1.19) |
| Diagnosis Code: B3 (Other Viral Diseases; Mycoses) | 0.98 (0.87-1.1) |
| Diagnosis Code: B9 (Sequelae of Infectious and Parasitic Diseases; Bacterial, Viral, and other Infectious Agents) | 1.07 (0.9-1.28) |
| Diagnosis Code: E8 (Metabolic Disorders) | 0.86 (0.63-1.15) |
| Diagnosis Code: G4 (Episodic and Paroxysmal Disorders) | 1.04 (0.84-1.28) |
| Diagnosis Code: G8 (Cerebral Palsy and other Paralytic Syndromes) | 0.66 (0.43-0.94) |
| Diagnosis Code: H5 (Disorders of Ocular Muscles, Binocular Movement, Accommodation and Refraction; Visual Disturbances and Blindness; Other Disorders of Eye and Adnexa) | 1 (0.8-1.26) |
| Diagnosis Code: H6 (Diseases of External Ear; Diseases of Middle Ear and Mastoid) | 0.89 (0.71-1.11) |
| Diagnosis Code: H9 (Other Disorders of Ear) | 0.84 (0.62-1.14) |
| Diagnosis Code: J0 (Acute Upper Respiratory Infections) | 1.06 (0.95-1.18) |
| Diagnosis Code: J1 (Influenza and Pneumonia) | 0.83 (0.65-1.05) |
| Diagnosis Code: J2 (Other Acute Lower Respiratory Infections) | 0.95 (0.82-1.1) |
| Diagnosis Code: J3 (Other Diseases of Upper Respiratory Tract) | 1.1 (0.85-1.39) |
| Diagnosis Code: J4 (Chronic Lower Respiratory Diseases) | 1.02 (0.87-1.18) |
| Diagnosis Code: K0 (Diseases of Oral Cavity, Salivary Glands, and Jaws) | 0.99 (0.7-1.43) |
| Diagnosis Code: K2 (Diseases of Oesophagus, Stomach, and Duodenum) | 1.01 (0.78-1.29) |
| Diagnosis Code: K5 (Noninfective Enteritis and Colitis; Other Diseases of Intestines) | 1.02 (0.88-1.19) |
| Diagnosis Code: L0 (Infections of the Skin and Subcutaneous Tissue) | 1.08 (0.84-1.36) |
| Diagnosis Code: L3 (Dermatitis and Eczema) | 1.06 (0.76-1.39) |
| Diagnosis Code: M2 (Arthropathies) | 1.25 (0.95-1.61) |
| Diagnosis Code:N3 (Other Diseases of Urinary System) | 0.99 (0.79-1.23) |
| Diagnosis Code: N4 (Diseases of Male Genital Organs) | 0.97 (0.76-1.22) |
| Diagnosis Code: P0 (Fetus and Newborn Affected by Maternal Factors and by Complications of Pregnancy, Labour, and Delivery; Disorders Related to Gestation and Fetal Growth) | 1.16 (0.91-1.45) |
| Diagnosis Code: P2 (Respiratory and Cardiovascular Disorders Specific to the | 1.17 (0.95-1.44) |

| | |
|---|---|
| Perinatal Period) | |
| Diagnosis Code: P3 (Infections Specific to the Perinatal Period) | 0.92 (0.69-1.22) |
| Diagnosis Code: P5 (Haemorrhagic and Haematological Disorders Specific to Fetus and Newborn) | 0.73 (0.55-0.95) |
| Diagnosis Code: P7 (Transitory Endocrine and Metabolic Disorders Specific to Fetus and Newborn; Conditions Involving the Integument and Temperature Regulation of Fetus and Newborn) | 0.89 (0.67-1.18) |
| Diagnosis Code: P9 (Other Disorders Originating in the Perinatal Period) | 1.01 (0.79-1.25) |
| Diagnosis Code: Q2 (Congenital Malformations of the Circulatory System) | 0.79 (0.58-1.05) |
| Diagnosis Code: Q5 (Congenital Malformations of Genital Organs) | 1.13 (0.84-1.52) |
| Diagnosis Code: Q6 (Congenital Malformations of the Urinary System; Congenital Malformations and Deformations of the Musculoskeletal System) | 0.91 (0.67-1.2) |
| Diagnosis Code: R0 (Symptoms and Signs Involving the Circulatory and Respiratory Systems) | 1.08 (0.94-1.23) |
| Diagnosis Code: R1 (Symptoms and Signs Involving the Digestive System and Abdomen) | 1.04 (0.91-1.19) |
| Diagnosis Code: R2 (Symptoms and Signs Involving the Nervous and Musculoskeletal Systems) | 1.04 (0.88-1.22) |
| Diagnosis Code: R4 (Symptoms and Signs Involving Cognition, Perception, Emotional State and Behaviour) | 2.09 (1.74-2.51) |
| Diagnosis Code: R5 (General Symptoms and Signs) | 1.02 (0.89-1.17) |
| Diagnosis Code: R6 (General Symptoms and Signs) | 0.92 (0.78-1.09) |
| Diagnosis Code: S0 (Injuries to the Head) | 1.03 (0.86-1.25) |
| Diagnosis Code: S5 (Injuries to the Elbow and Forearm) | 1.38 (1.08-1.74) |
| Diagnosis Code: S6 (Injuries to the Wrist and Hand) | 1.33 (1-1.72) |
| Diagnosis Code: S8 (Injuries to the Knee and Lower Leg) | 0.95 (0.73-1.24) |
| Diagnosis Code: T1 (Injuries to Unspecified Part of Trunk, Limb, or Body Region; Effects of Foreign Body Entering Through Natural Orifice) | 1.11 (0.83-1.44) |
| Diagnosis Code: T3 (Burns and Corrosions; Frostbite; Poisoning by Drugs, Medicaments and Biological Substances) | 1.07 (0.85-1.36) |
| Diagnosis Code: T4 (Poisoning by Drugs, Medicaments and Biological Substances) | 1.2 (0.91-1.56) |
| Diagnosis Code: T8 (Complications of Surgical and Medical Care, Not Elsewhere Classified) | 0.78 (0.57-1.09) |
| Diagnosis Code: U5 (Provisional Assignment of New Diseases of Uncertain Etiology or Emergency Use) | 1.22 (0.99-1.49) |
| Diagnosis Code: W0 (Falls) | 0.96 (0.78-1.2) |

| | |
|---|---|
| Diagnosis Code: W1 (Falls) | 1.04 (0.83-1.29) |
| Diagnosis Code: W2 (Exposure to Inanimate Mechanical Forces) | 0.91 (0.69-1.18) |
| Diagnosis Code: X4 (Accidental Poisoning by and Exposure to Noxious Substances ) | 1.05 (0.8-1.34) |
| Diagnosis Code: X5 (Overexertion, Travel and Privation; Accidental Exposure to Other and Unspecified Factors) | 0.87 (0.65-1.11) |
| Diagnosis Code: X6 (Intentional Self-Harm) | 2.8 (2.01-3.84) |
| Diagnosis Code: Z0 (Persons Encountering Health Services for Examination and Investigation) | 1.2 (1.02-1.42) |
| Diagnosis Code: Z3 (Persons Encountering Health Services in Circumstances Related to Reproduction) | 1.06 (0.91-1.23) |
| Diagnosis Code: Z4 (Persons Encountering Health Services for Specific Procedures and Health Care) | 1.01 (0.77-1.33) |
| Diagnosis Code: Z5 (Persons Encountering Health Services for Specific Procedures and Health Care; Persons with Potential Health Hazards Related to Socioeconomic and Psychosocial Circumstances) | 0.94 (0.77-1.14) |
| Diagnosis Code: Z6 (Persons with Potential Health Hazards Related to Socioeconomic and Psychosocial Circumstances) | 1.18 (0.99-1.43) |
| Diagnosis Code: Z7 (Persons Encountering Health Services in Other Circumstances ) | 1.11 (0.9-1.38) |
| Diagnosis Code: Z8 (Persons with Potential Health Hazards Related to Family and Personal History and Certain Conditions Influencing Health Status) | 0.9 (0.76-1.04) |
| Diagnosis Code: Z9 (Persons with Potential Health Hazards Related to Family and Personal History and Certain Conditions Influencing Health Status) | 1.51 (1.28-1.81) |
| Disability (Memory): Has Disability | 1.17 (1-1.4) |
| Disability (Mobility): Has Disability | 0.79 (0.63-0.96) |
| Disability: Has Disability | 1.44 (1.2-1.75) |
| Disability (Sensory): Has Disability | 0.64 (0.53-0.75) |
| Ethnicity: Asian | 0.41 (0.28-0.57) |
| Ethnicity: Black, African, Caribbean or Black British | 0.56 (0.36-0.85) |
| Ethnicity: Mixed Ethnic Groups | 0.82 (0.65-1.04) |
| Ethnicity: Information not Obtained | 0.92 (0.73-1.16) |
| Ethnicity: Other Ethnic Group | 0.69 (0.41-1.11) |
| Ethnicity: Information Refused | 0.73 (0.42-1.16) |
| School Exclusion Category: Fixed Term Exclusion | 1.72 (1.56-1.91) |
| School Exclusion Category: Permanent Exclusion | 1.63 (1.17-2.23) |

| | |
|---|---|
| Free School Meal Status: Eligible | 0.91 (0.82-1) |
| Free School Meal Status: Unknown | 0.95 (0.49-1.67) |
| Gender: Female | 0.78 (0.72-0.85) |
| Gestation Age: Continuous Value | 1.01 (0.94-1.07) |
| Autistic Spectrum Disorder Status: Autistic | 1.84 (1.6-2.13) |
| Autistic Spectrum Disorder Status: Unknown | 0.85 (0.62-1) |
| Dental Check Status: Not Receiving Checks | 0.82 (0.74-0.9) |
| Dental Check Status: Unknown | 0.88 (0.71-1.08) |
| Immunisation Status: Not Immunised | 0.78 (0.69-0.87) |
| Immunisation Status: Unknown | 0.99 (0.75-1.33) |
| Substance Misuse: Misusing Substances | 3.03 (2.68-3.46) |
| Substance Misuse: Unknown | 1.59 (0.57-4.15) |
| Labour Onset: Caesarean Section | 1.05 (0.88-1.23) |
| Labour Onset: Surgical Induction (amniotomy) | 1.51 (1.21-1.88) |
| Labour Onset: Medical Induction | 1.02 (0.57-1.75) |
| Labour Onset: Onset Not Known | 0.93 (0.52-1.57) |
| Labour Onset: Unknown | 1.19 (1.01-1.38) |
| Looked After Child Status: Looked After | 1.8 (1.63-1.98) |
| Maternal Smoking: Gave up during pregnancy | 0.84 (0.53-1.32) |
| Maternal Smoking: 0-9 cigarettes per day | 0.93 (0.74-1.14) |
| Maternal Smoking: 10-19 cigarettes per day | 1.03 (0.85-1.24) |
| Maternal Smoking: 20-29 cigarettes per day | 0.5 (0.13-1.28) |
| Maternal Smoking: Non-smoker | 0.9 (0.68-1.17) |
| Maternal Smoking: Unknown | 1.08 (0.92-1.31) |
| Apgar 1-Minute Score: Unknown | 1.03 (0.9-1.19) |
| Apgar 5-Minute Score: Unknown | 1.33 (1.16-1.53) |
| Birth Weight: Unknown | 0.75 (0.57-0.99) |
| Gestation Age: Unknown | 1.26 (0.99-1.62) |
| Welsh Index of Multiple Deprivation: Unknown | 0.31 (0.14-0.68) |
| Category of Need: Abuse or Neglect | 0.92 (0.82-1.03) |
| Category of Need: Child’s Disability or Illness | 1.24 (1.04-1.49) |
| Category of Need: Parental Disability or Illness | 1.1 (0.87-1.35) |
| Category of Need: Family in Acute Stress | 1.44 (1.26-1.64) |

| | |
|---|---|
| Category of Need: Family Dysfunction | 1.13 (1-1.28) |
| Category of Need: Socially Unacceptable Behaviour | 1.5 (1.25-1.79) |
| Category of Need: Low Income | 0.84 (0.25-2.16) |
| Category of Need: Absent Parenting | 0.86 (0.61-1.17) |
| Category of Need: Adoption Disruption | 1.95 (0.91-3.67) |
| Operation Code: A5 (Other operations on meninges of spinal cord; Therapeutic epidural injection; Drainage of spinal canal; Therapeutic spinal puncture; Diagnostic spinal puncture; Operations on spinal nerve root; Excision of peripheral nerve) | 1.13 (0.83-1.5) |
| Operation Code: D1 (Exenteration of mastoid air cells; Other operations on mastoid; Attachment of bone anchored hearing prosthesis; Repair of eardrum; Drainage of middle ear; Reconstruction of ossicular chain; Other operations on ossicle of ear; Extirpation of lesion of middle ear) | 1.13 (0.86-1.48) |
| Operation Code: E2 (Operations on adenoid; Repair of pharynx; Other open operations on pharynx; Therapeutic endoscopic operations on pharynx; Diagnostic endoscopic examination of pharynx; Other operations on pharynx; Operations on cricopharyngeus muscle; Excision of larynx) | 1.07 (0.8-1.45) |
| Operation Code: F1 (Simple extraction of tooth; Preprosthetic oral surgery; Surgery on apex of tooth; Restoration of tooth; Orthodontic operations; Other orthodontic operations; Other operations on tooth; Operations on teeth using dental crown or bridge; Excision of dental lesion of jaw) | 1.18 (0.83-1.7) |
| Operation Code: F3 (Other repair of palate; Other operations on palate; Excision of tonsil; Other operations on tonsil; Extirpation of lesion of other part of mouth; Reconstruction of other part of mouth) | 0.94 (0.74-1.22) |
| Operation Code: G4 (Incision of pylorus; Other operations on pylorus; Other fibreoptic endoscopic extirpation of lesion of upper gastrointestinal tract; Fibreoptic endoscopic extirpation of lesion of upper gastrointestinal tract; Other therapeutic fibreoptic endoscopic operations on upper gastrointestinal tract; Diagnostic fibreoptic endoscopic examination of upper gastrointestinal tract; Therapeutic fibreoptic endoscopic operations on upper gastrointestinal tract; Intubation of stomach; Other operations on stomach; Excision of duodenum.) | 0.88 (0.63-1.2) |
| Operation Code: S4 (Other closure of skin; Suture of skin of head or neck; Suture of skin of other site; Removal of repair material from skin; Removal of other inorganic substance from skin; Removal of other substance from skin; Opening of skin; Insertion of skin expander into subcutaneous tissue; Attention to skin expander in subcutaneous tissue) | 1.1 (0.81-1.46) |
| Operation Code: S5 (Introduction of other inert substance into subcutaneous tissue; Introduction of destructive substance into subcutaneous tissue; Introduction of therapeutic substance into subcutaneous tissue, Introduction of substance into skin; Exploration of burnt skin of head or neck; Exploration of burnt | 1.16 (0.86-1.54) |

| | |
|---|---|
| skin of other site; Exploration of other skin of head or neck; Exploration of other skin of other site; Larvae therapy of skin; Leech therapy of skin) | |
| Operation Code: U0 (Diagnostic imaging of whole body; Diagnostic imaging of mouth; Diagnostic imaging of central nervous system; Diagnostic imaging of face and neck; Diagnostic imaging of chest; Diagnostic imaging of abdomen; Diagnostic imaging of pelvis) | 0.67 (0.5-0.92) |
| Operation Code: U2 (Diagnostic echocardiography; Diagnostic imaging procedures; Neuropsychology tests; Nuclear medicine haematological tests, Diagnostic audiology, Breath tests, Diagnostic testing of genitourinary system; Diagnostic application tests on skin; Other diagnostic tests on skin, Diagnostic endocrinology) | 0.8 (0.57-1.05) |
| Operation Code: W2 (Primary open reduction of fracture of bone and extramedullary fixation; Primary open reduction of intra-articular fracture of bone; Other primary open reduction of fracture of bone; Secondary open reduction of fracture of bone; Closed reduction of fracture of bone and internal fixation; Closed reduction of fracture of bone and external fixation; Other closed reduction of fracture of bone; Fixation of epiphysis; Other internal fixation of bone; Skeletal traction of bone) | 1.28 (0.86-1.78) |
| Operation Code: X2 (Correction of congenital deformity of forearm; Correction of congenital deformity of hand; Correction of congenital deformity of hip; Correction of congenital deformity of leg; Primary correction of congenital deformity of foot; Other correction of congenital deformity of foot; Correction of minor congenital deformity of foot; Intermittent infusion of therapeutic substance; Continuous Infusion of therapeutic substance) | 1.09 (0.81-1.41) |
| Operation Code: X3 (Injection of therapeutic substance; Injection of radiocontrast material; Exchange blood transfusion; Other blood transfusion; Other intravenous transfusion; Other intravenous injection; Blood withdrawal; Intramuscular injection; Subcutaneous injection; Other route of administration of therapeutic substance) | 0.81 (0.63-1.02) |
| Operation Code: X5 (External resuscitation; Change of body temperature; Oxygen therapy; Extirpation of unspecified organ; Other operations on unspecified organ; Intubation of trachea; Artificial support for body system; Anaesthetic without surgery) | 1.04 (0.76-1.43) |
| Operation Code: Y5 (Approach through abdominal cavity; Approach to organ through artificial opening into gastrointestinal tract; Approach to organ through other opening; Approach to organ under image control; Harvest of nerve; Harvest of random pattern flap of skin from limb; Harvest of random pattern flap of skin from other site; Harvest of axial pattern flap of skin; Harvest of skin for graft; Harvest of flap of skin and fascia) | 0.84 (0.63-1.12) |
| Operation Code: Y7 (Early operations NOC; Late operations NOC; Facilitating operations NOC; Minimal access to thoracic cavity; Minimal access to abdominal cavity; Minimal access to other body cavity; Arteriotomy approach to organ under | 0.91 (0.67-1.25) |

| | |
|---|---|
| image control; Approach to organ through artery) | |
| Operation Code: Y8 (General Anaesthetic; Spinal Anaesthetic; Local Anaesthetic; Other Anaesthetic; Y89 Brachytherapy) | 1.03 (0.89-1.16) |
| Operation Code: Y9 (Other non-operations; External beam radiotherapy; Support for preparation for radiotherapy; Gallium-67 imaging; Radiopharmaceutical imaging; Gestational age; In vitro fertilisation; Radiology with contrast; Y98 Radiology procedures; Y99 Donor status) | 0.97 (0.73-1.27) |
| Operation Code: Z4 (Other vascular tissue; Upper urinary tract; Lower urinary tract; Male genital organ; Vagina; Uterus; Other female genital tract; Skin of face; Skin of other part of head or neck; Skin of trunk) | 1.02 (0.69-1.48) |
| Operation Code: Z5 (Skin of other site; Nail; Chest wall; Abdominal wall; Muscle of shoulder or upper arm; Muscle of forearm; Muscle of hand; Muscle of hip or thigh; Muscle of lower leg; Muscle of foot) | 0.94 (0.72-1.25) |
| Operation Code: Z7 (Radius; Ulna; Other bone of arm or wrist; Other bone of hand; Rib cage; Bone of pelvis; Femur; Tibia; Bone of tarsus) | 0.68 (0.48-0.99) |
| Operation Code: Z8 (Other bone of foot; Joint of shoulder girdle or arm; Joint of wrist or hand; Joint of finger; Joint of pelvis or upper leg; Joint of lower leg or tarsus; Other joint of foot; Other part of musculoskeletal system; Respiratory tract; Arm region) | 0.94 (0.7-1.21) |
| Operation Code: Z9 (Leg region; Other vein of upper body; Other region of body; Other veins of pelvis; Laterality of operation; Other branch of thoracic aorta; Other lateral branch of abdominal aorta; Other terminal branch of aorta; Other veins of lower limb; Intervertebral disc) | 0.87 (0.75-1.01) |
| Parenting Capacity (Domestic Abuse): Abuse | 0.82 (0.74-0.91) |
| Parenting Capacity (Domestic Abuse): Unknown | 1.72 (0.52-4.67) |
| Parenting Capacity (Learning Disabilities): Learning Disabilities | 1.03 (0.88-1.18) |
| Parenting Capacity (Learning Disabilities): Unknown | 1.53 (0.66-3.73) |
| Parenting Capacity (Mental Health): Mental Health Issues | 1.54 (1.39-1.68) |
| Parenting Capacity (Mental Health): Unknown | 0.33 (0.15-0.84) |
| Parenting Capacity (Physical Health): Physical Health Issues | 1.09 (0.99-1.19) |
| Parenting Capacity (Physical Health): Unknown | 1.03 (0.46-2.52) |
| Parenting Capacity (Substance Misuse): Substance Misuse | 0.78 (0.7-0.87) |
| Parenting Capacity (Substance Misuse): Unknown | 0.53 (0.26-1.26) |
| Welsh Index of Multiple Deprivation: Continuous Value | 1.06 (1.01-1.11) |
| Youth Offending Status: Offender | 1.24 (1.05-1.45) |
| Youth Offending Status: Unknown | 1.83 (0.74-4.03) |

**Table 5a: Fairness Metrics (SVM Model)**

| | TPR | TNR | PPV | NPV |
|---|---|---|---|---|
| **Asian** | 0.091 | 1.0 | 1.0 | 0.861 |
| **Black** | 0.0 | 1.0 | 0.0 | 0.918 |
| **Mixed** | 0.154 | 9.61 | 0.400 | 0.871 |
| **Other** | 0.400 | 1.0 | 1.0 | 0.938 |
| **White** | 0.174 | 0.973 | 0.616 | 0.826 |
| **Female** | 0.227 | 0.971 | 0.692 | 0.816 |
| **Male** | 0.105 | 0.977 | 0.481 | 0.841 |

**Table 5b: Fairness Metrics (Logistic Regression Model)**

| | TPR | TNR | PPV | NPV |
|---|---|---|---|---|
| **Asian** | 0.364 | 0.935 | 0.5 | 0.892 |
| **Black** | 0.0 | 0.933 | 0.0 | 0.913 |
| **Mixed** | 0.538 | 0.688 | 0.226 | 0.898 |
| **Other** | 1.0 | 0.844 | 0.417 | 1.0 |
| **White** | 0.592 | 0.700 | 0.330 | 0.873 |
| **Female** | 0.599 | 0.689 | 0.352 | 0.859 |
| **Male** | 0.560 | 0.731 | 0.301 | 0.889 |
